# Health system and epidemiological determinants of mortality in acute lymphoblastic leukemia in the Brazilian Amazon

**DOI:** 10.64898/2026.02.23.26346933

**Authors:** Isabelle Vasconcelos Sousa, Fábio Magalhães-Gama, Beatriz Silva Oliveira, Estela Yasmin Coelho Oliveira, Julia Goes Souza Ghedini, Leticia Paes Araujo Carvalho, Joey Ramone Ferreira Fonseca, Vitória Giovanna Rodrigues Santos, Juniel Assis Crespo-Neto, Flávia Santos Pio, Julia Santos Moraes, Mateus Souza Barros, Flávio Souza Silva, Claudio Lucas Santos Catão, Maria Gabriela Almeida Rodrigues, Sheila Vitor-Silva, Fabíola Silva Alves-Hanna, Allyson Guimarães Costa

**Author notes:** Corresponding Author: Allyson Guimarães Costa, UFAM, Av. Rodrigo Otávio Jordão Ramos, 1200, Coroado, Manaus, AM, CEP 69067-005, Brazil.

## Abstract

**Background:** Acute lymphoblastic leukemia is the most common childhood malignancy; however, survival remains highly heterogeneous across regions, particularly in low- and middle-income settings. In geographically vulnerable areas such as the Brazilian Amazon, structural barriers related to health system organization, centralization of specialized services, and continuity of care may substantially influence treatment trajectories and mortality. This study aimed to examine health system and epidemiological determinants of mortality among patients with acute lymphoblastic leukemia in Amazonas, Brazil.

**Methods:** This retrospective cohort study included 393 patients diagnosed with acute lymphoblastic leukemia between 2016 and 2021 at the state referral center for hematologic diseases in Amazonas, Brazil. Sociodemographic, clinical, laboratory, and health system–related variables were analyzed. Multivariate logistic regression models were used to identify factors associated with mortality. Spatial analyses described the geographic distribution of cases, and overall survival was evaluated using Kaplan-Meier curves.

**Results:** The cohort was predominantly pediatric, with a substantial proportion of adolescents and young adults. Most patients presented with high-risk clinical and laboratory features at diagnosis. Mortality occurred in 48.5% of patients and was strongly associated with age at diagnosis, with higher odds of death among adolescents, young adults, and individuals aged 51–60 years. Geographic concentration of specialized services and treatment-related trajectories were closely linked to survival patterns.

**Conclusion:** Mortality from acute lymphoblastic leukemia in the Brazilian Amazon remains high and is primarily influenced by age-related vulnerability and health system factors rather than baseline sociodemographic characteristics. These findings underscore persistent regional inequalities in access to specialized care and highlight the need for health system strengthening to improve cancer survival in geographically vulnerable settings.

## INTRODUCTION

Acute lymphoblastic leukemia (ALL) is the most common malignant neoplasm in childhood, accounting for approximately 75% of pediatric leukemias, with an estimated incidence ranging from 1.5 to 4.5 cases per 100,000 individuals per year worldwide (1). Over recent decades, advances in risk stratification, chemotherapeutic regimens, and supportive care have enabled high-income countries to achieve survival rates exceeding 85% among pediatric patients (2). However, these gains have not been equitably distributed. In many low-and middle-income countries (LMICs), survival remains substantially lower, reflecting persistent disparities in timely diagnosis, access to specialized treatment, continuity of care, and supportive management (3). These global survival gaps highlight the central role of health system capacity and organization in shaping cancer outcomes.

Beyond biological characteristics, demographic and clinical factors contribute to outcome variability in ALL. Age at diagnosis is one of the most consistent prognostic determinants, with poorer survival observed among adolescents, young adults, and older patients compared with younger children (4). Nevertheless, in resource-constrained settings, clinical vulnerability often interacts with structural determinants, including delayed referral, fragmented care pathways, and limited availability of high-complexity services, amplifying mortality risks (5). Understanding how epidemiological and health system factors jointly influence survival is therefore essential to addressing inequities in childhood and adolescent cancer outcomes globally.

The Brazilian Amazon represents a geographically and socially vulnerable region within a middle-income country. The state of Amazonas is characterized by vast territorial extension, low population density, reliance on river transportation, and marked centralization of high-complexity health services in the capital city (6,7). These structural features create barriers to timely diagnosis, treatment initiation, and continuity of care for patients residing in remote municipalities. In such contexts, mortality from potentially curable diseases such as ALL may reflect not only disease severity but also systemic limitations in service organization and accessibility.

Previous regional analyses conducted at the Fundação Hospitalar de Hematologia e Hemoterapia do Amazonas (HEMOAM) between 2005 and 2015 described the epidemiological profile of acute leukemias and identified factors associated with comorbidities and mortality (8). However, updated evidence is needed to assess whether health system–related challenges continue to shape survival patterns and to better understand how age-related vulnerability and geographic barriers interact with mortality in this setting.

## METHODS

### Ethics

Ethical approval for this study was obtained from the Ethics Committee of the Fundação Hospitalar de Hematologia e Hemoterapia do Amazonas (approval number 4.982.395). Further details are provided in the Declarations section.

### Study Setting

The study was conducted in the state of Amazonas, Brazil, a geographically vast and sparsely populated region in the Brazilian Amazon. According to the Brazilian Institute of Geography and Statistics (IBGE), Amazonas covers 1,559,146.876 km² and comprises 62 municipalities. The region is characterized by extensive geographic dispersion, low population density, and reliance on river transportation, factors that substantially affect access to specialized healthcare services (7,9).

High-complexity medical services are largely centralized in Manaus, the state capital, which concentrates healthcare infrastructure and referral centers. The Fundação Hospitalar de Hematologia e Hemoterapia do Amazonas (HEMOAM), located in Manaus, functions as the state referral center for diagnosis, treatment, and follow-up of hematologic malignancies. It receives patients from remote riverside and inland municipalities across the state and occasionally from neighboring northern states (7,8).

### Study Population and Design

This retrospective cohort study included all patients diagnosed with acute lymphoblastic leukemia (ALL) between January 2016 and December 2021 who received treatment and follow-up at HEMOAM. Cases were classified according to the International Classification of Diseases, 10th Revision (ICD-10 code C91), and diagnosis was confirmed by immunophenotyping.

### Eligibility Criteria

Patients were eligible if they had a confirmed diagnosis of ALL and received treatment at HEMOAM during the study period. Patients with inconclusive diagnostic records or other hematologic malignancies were excluded. Cases were also excluded if medical records were unavailable or duplicate entries were identified. A total of 393 patients met the eligibility criteria and were included in the final analysis.

### Data Sources and Variables

Data were obtained from institutional medical records and laboratory information systems to ensure completeness and minimize information loss. Primary data were obtained from the Medical and Statistical Care System (SAME), including both physical medical records archived in the Patient Records Service (SSP) and electronic records accessed through the iDoctor^®^ system. Additional information was retrieved from social service files and laboratory records available in the SoftLab^®^ system, allowing the recovery of sociodemographic, clinical, and laboratory data not consistently documented across individual medical charts. Sociodemographic variables included age at diagnosis, sex, race/ethnicity, schooling level, per capita household income, and municipality of residence. These variables were used to assess potential epidemiological and structural determinants of mortality.

Clinical variables included immunophenotypic profile, cytogenetic risk features, laboratory parameters at diagnosis, and treatment characteristics. Mortality from any cause during follow-up was defined as the primary outcome and was ascertained through medical records, institutional surveillance systems, and linkage with the Brazilian National Death Registry.

### Statistical Analysis

Descriptive statistics were used to summarize baseline characteristics. Multivariate logistic regression models were applied to identify epidemiological and health system–related factors associated with mortality. Covariates included age at diagnosis, sex, race/ethnicity, schooling level, family income, municipality of residence, and clinical risk indicators. Results were expressed as odds ratios (ORs) with 95% confidence intervals (95% CIs).

Overall survival was estimated using Kaplan–Meier curves, and survival differences were assessed using the log-rank test. Spatial distribution of cases across municipalities was mapped using QGIS software to explore geographic patterns in access and outcomes. Statistical analyses were conducted using STATA version 13 (StataCorp, College Station, TX, USA).

## RESULTS

### Epidemiological and Sociodemographic Profile

A total of 393 patients with confirmed acute lymphoblastic leukemia (ALL) were included. The cohort was predominantly pediatric, with 52.8% aged 0–10 years and 21.4% aged 11–20 years. Only 6.1% were older than 50 years at diagnosis. Males accounted for 58.3% of cases (**Table 1**).

**Table 1.** Epidemiological and sociodemographic characteristics of patients with acute lymphoblastic leukemia in the state of Amazonas, Brazil, 2016–2021.

| Variables | n (%) |
| --- | --- |
| Age in Years |  |
| 0–10 | 207/393 (52.77) |
| 11–20 | 84/393 (21.37) |
| 21–30 | 35/393 (8.91) |
| 31–40 | 22/393 (5.60) |
| 41–50 | 21/393 (5.34) |
| 51–60 | 12/393 (3.05) |
| >60 | 12/393 (3.05) |
| **Sex** |  |
| Male | 229/393 (58.27) |
| Female | 164/393 (41.73) |
| **Race/ethnicity** |  |
| White | 24/279 (8.60) |
| Mixed ethnicity | 226/279 (81.00) |
| Black | 4/279 (1.43) |
| Indigenous | 19/279 (6.81) |
| Asian | 6/279 (2.15) |
| **Marital status** |  |
| Single | 218/258 (84.50) |
| Married | 30/258 (11.63) |
| Divorced | 2/258 (0.78) |
| Widowed | 2/258 (0.78) |
| Consensual union / cohabitation | 6/258 (2.33) |
| **Schooling** |  |
| None | 3/86 (3.49) |
| 1–4 years | 51/86 (59.30) |
| 5–8 years | 4/86 (4.65) |
| ≥9 years | 28/86 (32.56) |
| **Monthly household income** |  |
| <1 minimum wage | 50/67 (74.63) |
| 1 minimum wage | 10/67 (14.93) |
| >1 minimum wage | 7/67 (10.45) |
| **Place of Residence** |  |
| Manaus | 196/360 (54.44) |
| Interior of Amazonas | 148/360 (41.11) |
| Other states/countries | 16/360 (4.44) |
| **Family history of cancer** | 33/393 (8.40) |

Values are presented as n (%). Proportions were calculated based on the total number of patients with available information for each variable.

Among patients with available race/ethnicity data, most self-identified as pardo (mixed ethnicity), reflecting the demographic composition of the region. Educational attainment and income data, although incomplete, indicated substantial social vulnerability, with the majority of recorded households reporting income below one minimum wage.

More than half of patients resided in Manaus, the state capital. However, 41.1% originated from municipalities in the interior of Amazonas, underscoring the regional dependence on centralized high-complexity services. In addition, a family history of cancer was reported in 8.4% of patients.

### Clinical Characteristics at Diagnosis

B-cell ALL was the predominant subtype (90.3%), while T-cell ALL accounted for 8.9% of cases. High-risk cytogenetic abnormalities were identified in 19.2% of patients with available data (**Table 2**).

**Table 2.** Clinical and laboratory characteristics of patients with acute lymphoblastic leukemia in the state of Amazonas, Brazil, 2016–2021.

| Variables | n (%) |
| --- | --- |
| **Immunophenotyping profile** |  |
| B-cell Acute Lymphoblastic Leukemia | 355/393 (90.33) |
| T-cell Acute Lymphoblastic Leukemia | 35/393 (8.91) |
| Mixed Phenotype Acute Leukemia | 3/393 (0.76) |
| **High-risk cytogenetic findings at diagnosis<sup>□</sup>** | 19/80 (19.19) |
| **Bone marrow blast percentage at diagnosis, median (%)<sup>‡</sup>** | 77.00 |
| **Central nervous system involvement at diagnosis** | 21/389 (5.40) |
| **Total leukocytes at diagnosis<sup>□</sup>** |  |
| < $15 \times 10^3/\mu\text{l}$ | 143/362 (60.50) |
| $\geq 15 \times 10^3/\mu\text{L}$ | 219/362 (39.50) |
| **Hemoglobin at diagnosis<sup>°</sup>** |  |
| < 10 g/dL | 303/362 (83.70) |
| $\geq 10$ g/dL | 59/362 (16.30) |
| **Platelets at diagnosis<sup>Δ</sup>** |  |
| < $100 \times 10^3/\mu\text{L}$ | 251/362 (69.34) |
| $\geq 100 \times 10^3/\mu\text{L}$ | 111/362 (30.66) |
| **Risk stratification at diagnosis** |  |
| Low risk | 77/328 (23.48) |
| Intermediate risk | 21/328 (6.40) |
| High risk | 230/328 (70.12) |
| **Treatment protocol** |  |
| GBTLI | 132/393 (33.59) |
| BFM | 128/393 (32.57) |
| GMALL | 87/393 (22.14) |
| Other protocols | 46/393 (11.70) |
| **Hematopoietic stem cell transplantation** | 12/393 (3.05) |
| **Transfusion history** | 273/374 (63.37) |
| **Non-infectious comorbidities** | 97/389 (24.94) |
| **Infectious comorbidities** | 181/389 (46.53) |
| **Mortality** | 183/377 (48.54) |

BFM: Berlin-Frankfurt-Münster; GBTLI: Grupo Brasileiro de Tratamento de Leucemia Infantil; GMALL: German Multicenter Study Group for Adult Acute Lymphoblastic Leukemia. Values are presented as n (%). Proportions were calculated based on the total number of patients with available information for each variable. <sup>□</sup>High-risk cytogenetic findings: translocation(9;22)(q34;q11), BCR-ABL1 Philadelphia chromosome; inversion(16)(p13;q22), myeloid co-expression. <sup>Δ</sup>Absolute lymphoblast count, calculated using the following formula: (Lymphoblast percentage from flow cytometry × total leukocyte count from hematological analyzer) / 100. <sup>°</sup>Total leukocyte count at diagnosis was categorized using a cutoff of $15 \times 10^3/\mu\text{L}$ , in accordance with established risk stratification criteria for acute lymphoblastic leukemia. <sup>°</sup>Hemoglobin level at diagnosis was categorized using a cutoff of 10 g/dL, reflecting clinically significant anemia commonly adopted in leukemia studies. <sup>Δ</sup>Platelet count at diagnosis was categorized using a cutoff of $100 \times 10^3/\mu\text{L}$ , consistent with standard definitions of thrombocytopenia.

Markers of advanced disease at presentation were frequent. Most patients exhibited elevated leukocyte counts, anemia, thrombocytopenia, and high bone marrow blast percentages at diagnosis. Risk stratification data showed that 70.1% of patients were classified as high risk.

During follow-up, infectious comorbidities were recorded in 46.5% of patients, and non-infectious comorbidities in 24.9%. Mortality occurred in 48.5% of patients with available outcome data.

### Spatial and Temporal Patterns

The spatial distribution of cases revealed marked geographic heterogeneity (**Figure 1**). While Manaus accounted for 54.4% of cases, a substantial proportion originated from inland municipalities, including Manacapuru, Tefé, and Careiro da Várzea.

**Figure 1.**
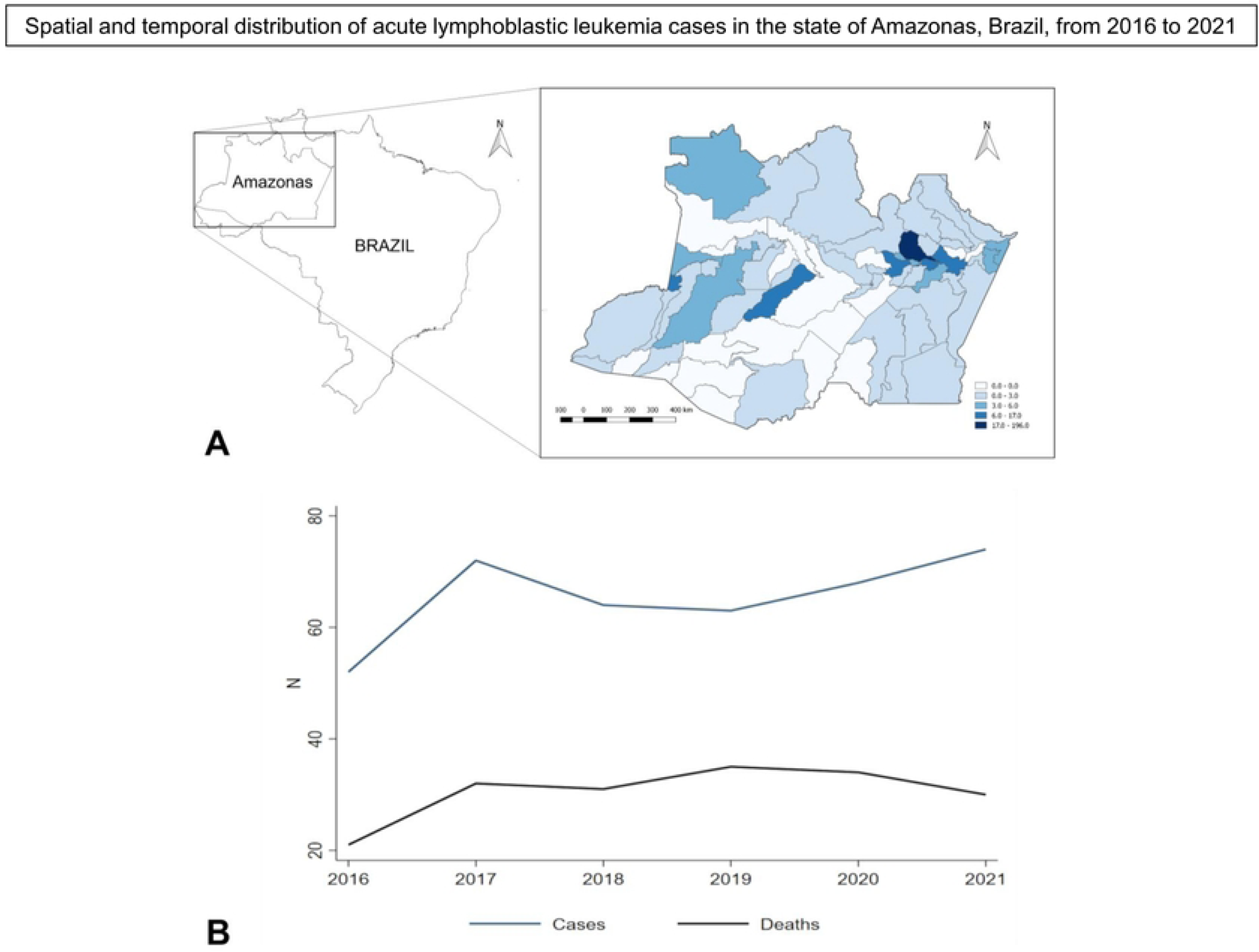
Spatial and temporal distribution of acute lymphoblastic leukemia cases in the state of Amazonas, Brazil, from 2016 to 2021. (A) Geographic distribution of acute lymphoblastic leukemia (ALL) cases across municipalities. Manaus, the state capital, accounted for the largest proportion of cases, while inland municipalities such as Manacapuru, Tefé, and Careiro da Várzea also showed notable case counts. A small number of patients originated from other states or neighboring countries (Colombia and Venezuela). (B) Annual number of ALL cases and deaths during the study period. Case counts fluctuated over time, with peaks in 2017 and 2019, and reached their highest level in 2021. Mortality peaked in 2019 before gradually declining through 2021.

Annual case counts fluctuated between 2016 and 2021, with peaks in 2017, 2019, and 2021. Mortality followed a similar pattern, with a modest peak in 2019 and gradual decline thereafter. These patterns indicate sustained regional burden and ongoing demand for specialized services.

### Determinants of Comorbidities

In multivariate analyses, sociodemographic variables were not independently associated with infectious comorbidities (**Table 3**). In contrast, non-infectious comorbidities were significantly associated with older age, particularly among patients aged 51–60 years (OR = 6.397; 95% CI: 1.543–26.517; p = 0.011) (**Supplementary Table S1**)

**Table 3.**
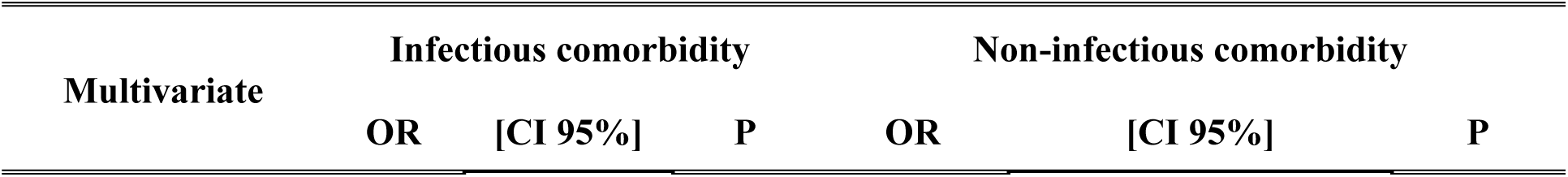
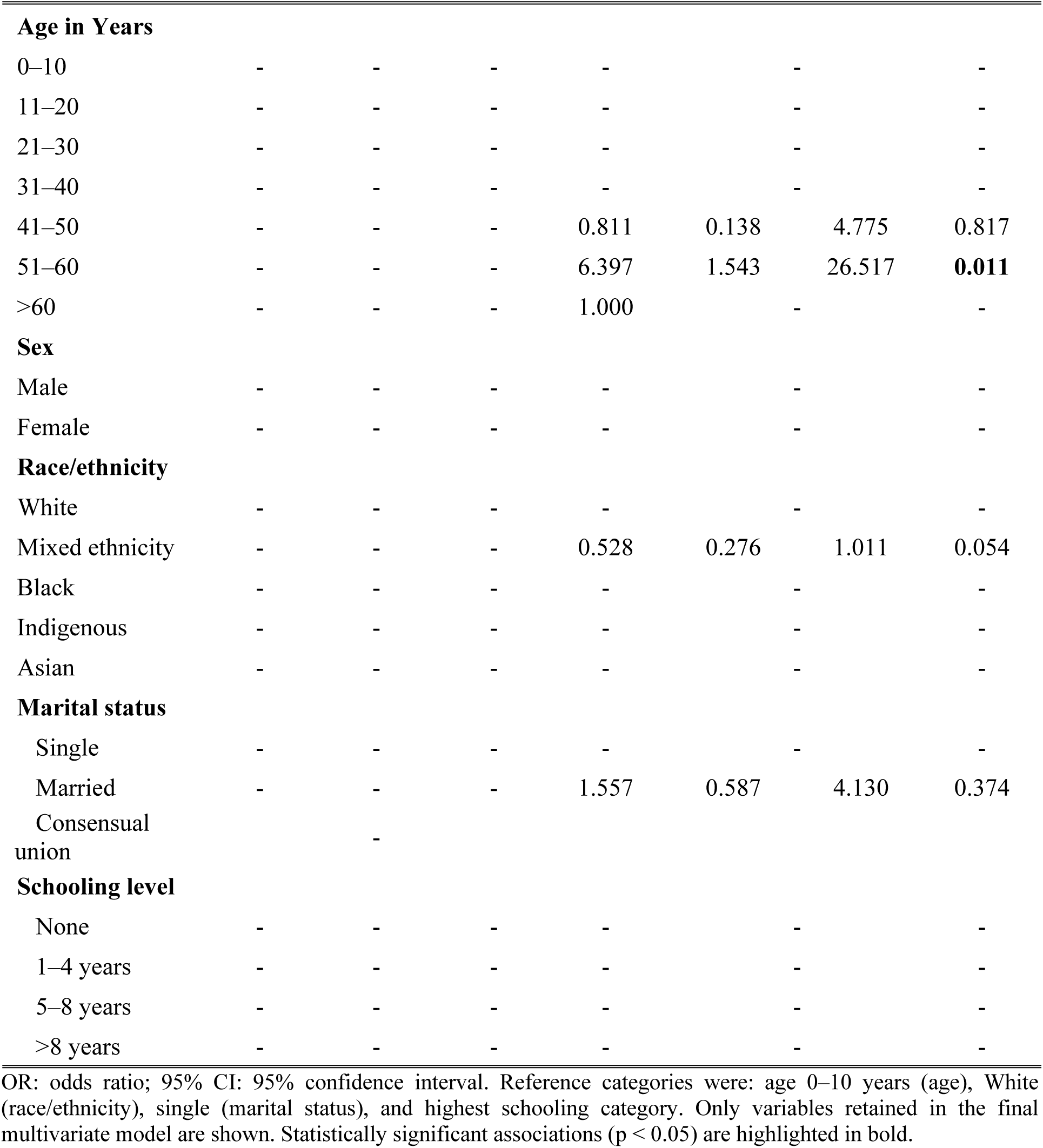
Multivariate logistic regression analysis of factors associated with infectious and non-infectious comorbidities among patients with acute lymphoblastic leukemia in Amazonas, Brazil, 2016–2021.

| Multivariate | Infectious comorbidity |  |  | Non-infectious comorbidity |  |  |
| --- | --- | --- | --- | --- | --- | --- |
|  | OR | [CI 95%] | P | OR | [CI 95%] | P |

|  |  |  |  |  |  |  |  |
| --- | --- | --- | --- | --- | --- | --- | --- |
| **Age in Years** |  |  |  |  |  |  |  |
| 0–10 | - | - | - | - | - | - | - |
| 11–20 | - | - | - | - | - | - | - |
| 21–30 | - | - | - | - | - | - | - |
| 31–40 | - | - | - | - | - | - | - |
| 41–50 | - | - | - | 0.811 | 0.138 | 4.775 | 0.817 |
| 51–60 | - | - | - | 6.397 | 1.543 | 26.517 | **0.011** |
| >60 | - | - | - | 1.000 | - | - | - |
| **Sex** |  |  |  |  |  |  |  |
| Male | - | - | - | - | - | - | - |
| Female | - | - | - | - | - | - | - |
| **Race/ethnicity** |  |  |  |  |  |  |  |
| White | - | - | - | - | - | - | - |
| Mixed ethnicity | - | - | - | 0.528 | 0.276 | 1.011 | 0.054 |
| Black | - | - | - | - | - | - | - |
| Indigenous | - | - | - | - | - | - | - |
| Asian | - | - | - | - | - | - | - |
| **Marital status** |  |  |  |  |  |  |  |
| Single | - | - | - | - | - | - | - |
| Married | - | - | - | 1.557 | 0.587 | 4.130 | 0.374 |
| Consensual union | - | - | - | - | - | - | - |
| **Schooling level** |  |  |  |  |  |  |  |
| None | - | - | - | - | - | - | - |
| 1–4 years | - | - | - | - | - | - | - |
| 5–8 years | - | - | - | - | - | - | - |
| >8 years | - | - | - | - | - | - | - |

OR: odds ratio; 95% CI: 95% confidence interval. Reference categories were: age 0–10 years (age), White (race/ethnicity), single (marital status), and highest schooling category. Only variables retained in the final multivariate model are shown. Statistically significant associations ( $p < 0.05$ ) are highlighted in bold.

### Determinants of Mortality

Age at diagnosis emerged as the strongest independent determinant of mortality. Compared with the reference group, patients aged 11–20 years (OR = 2.712; 95% CI: 1.305–5.634), 21–30 years (OR = 3.146; 95% CI: 1.178–8.400), and 51–60 years (OR = 7.952; 95% CI: 1.465–43.175) had significantly higher odds of death (**Table 4**).

**Table 4.** Multivariate logistic regression analysis of factors associated with mortality among patients with acute lymphoblastic leukemia in Amazonas, Brazil, from 2016 to 2021.

| Multivariate | Death |  |  |  |
| --- | --- | --- | --- | --- |
|  | OR | [CI 95%] |  | P |
| Age in Years |  |  |  |  |
| 0–10 | 1.000 | - |  | - |
| 11–20 | 2.712 | 1.305 | 5.634 | 0.008 |
| 21–30 | 3.146 | 1.178 | 8.400 | 0.022 |
| 31–40 | 1.998 | 0.413 | 9.660 | 0.389 |
| 41–50 | 1.000 | - |  | - |
| 51–60 | 7.952 | 1.465 | 43.175 | 0.016 |
| >60 | 0.939 | 0.050 | 17.685 | 0.967 |
| Sex |  |  |  |  |
| Male | - | - |  | - |
| Female | - | - |  | - |
| Race/ethnicity |  |  |  |  |
| White | - | - |  | - |
| Mixed ethnicity | - | - |  | - |
| Black | - | - |  | - |
| Indigenous | - | - |  | - |
| Asian | - | - |  | - |
| Marital status |  |  |  |  |
| Single | - | - |  | - |
| Married | 3.811 | 1.205 | 12.051 | 0.023 |
| Consensual union / cohabitation | - | - |  | - |
| Schooling |  |  |  |  |
| None | - | - |  | - |
| 1–4 years | - | - |  | - |
| 5–8 years | - | - |  | - |
| >8 years | - | - |  | - |
| Family history of cancer | - | - |  | - |
| **Non-infectious comorbidity** | 1.327 | 0.660 | 2.668 | 0.428 |
| **Infectious comorbidity** | 0.426 | 0.233 | 0.779 | **0.006** |

OR: odds ratio; 95% CI: 95% confidence interval. Reference categories were: age 0–10 years (age), White (race/ethnicity), single (marital status), highest schooling category, and absence of comorbidity or relapse. Only variables retained in the final multivariate model are shown. Statistically significant associations ( $p < 0.05$ ) are highlighted in bold.

Marital status was also associated with mortality, with married individuals exhibiting higher risk. No independent associations were observed for sex, race/ethnicity, schooling level, or family history of cancer. Infectious comorbidities were inversely associated with mortality (OR = 0.426; 95% CI: 0.233–0.779), whereas non-infectious comorbidities showed no significant association (**Supplementary Table S2**)

### Overall survival

Kaplan–Meier analysis demonstrated significantly longer overall survival among patients who developed infectious comorbidities during follow-up (log-rank p < 0.02). A total of 367 patients were included in the survival analysis, with 175 recorded deaths (**Figure 2**). No survival differences were observed according to non-infectious comorbidity status (**Supplementary Figure S1**).

**Figure 2.**
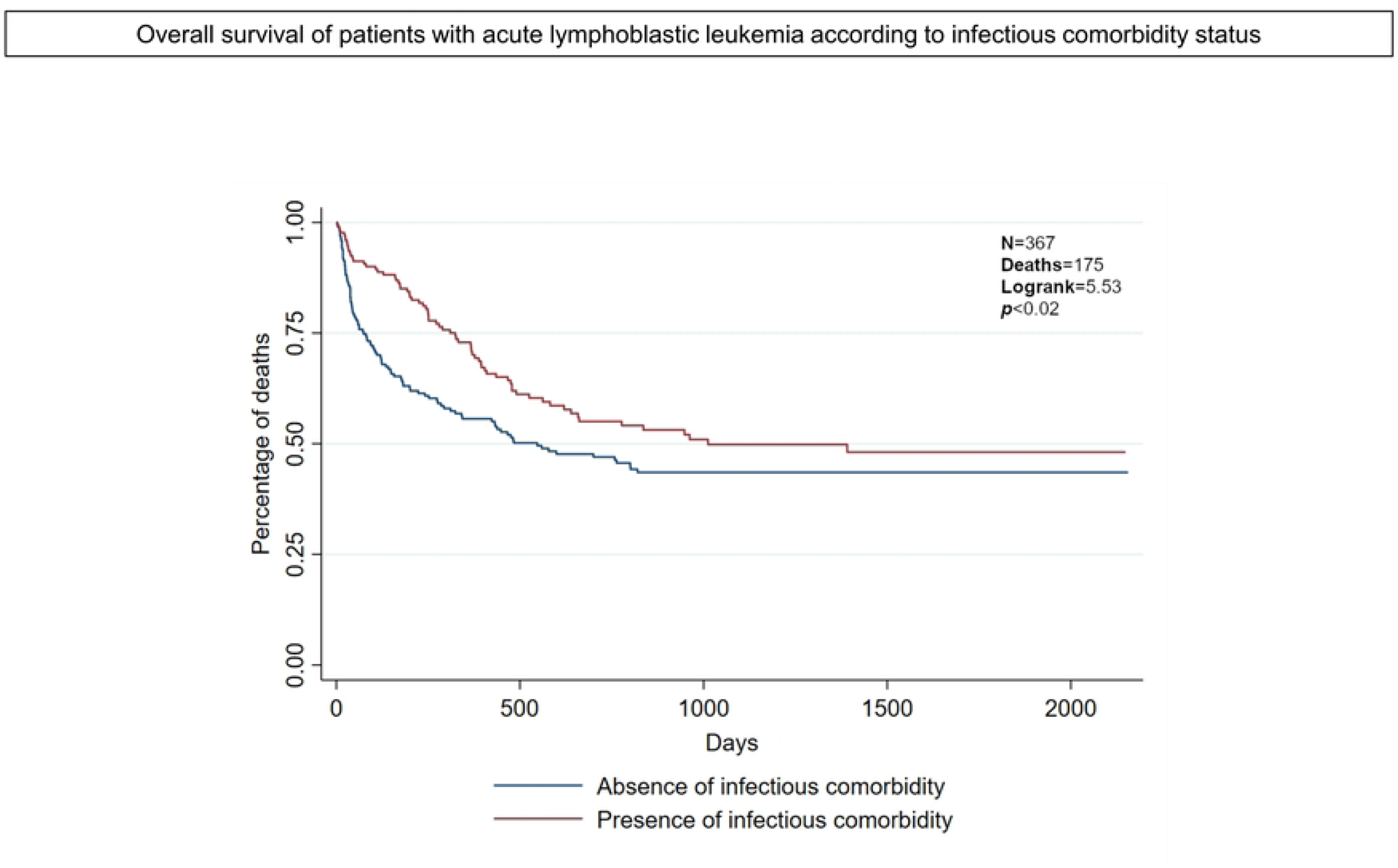
Overall survival of patients with acute lymphoblastic leukemia according to infectious comorbidity status. Kaplan–Meier curves depicting overall survival stratified by the presence or absence of infectious comorbidities during follow-up. The red line represents patients with infectious comorbidities, and the blue line represents patients without infectious comorbidities. Survival differences were assessed using the log-rank test (p < 0.02). A total of 367 patients were included in the analysis, with 175 deaths observed during follow-up.

## DISCUSSION

The epidemiological profile observed in this cohort reflects the expected predominance of pediatric cases of acute lymphoblastic leukemia (ALL); however, a substantial proportion of adolescents and young adults was also identified. This finding is particularly relevant given the consistently poorer outcomes reported outside the classical pediatric age range, reinforcing age at diagnosis as a central determinant of mortality in ALL, especially in low- and middle-income settings (1,2,10,11).

The sociodemographic characteristics of the study population further highlight a context of marked social vulnerability, characterized by low educational attainment, limited household income, and a high proportion of patients residing in municipalities outside the state capital. Although these variables were not independently associated with mortality in adjusted analyses, they provide important context for understanding disparities in access to care and treatment continuity, which are known to influence leukemia outcomes at the population level (6,8). In geographically vast regions such as Amazonas, structural disadvantages may operate indirectly through delayed diagnosis, referral pathways, and logistical barriers rather than appearing as isolated statistical predictors.

The geographic distribution of cases underscores the strong centralization of high-complexity hematologic care in Manaus, with more than 40% of patients originating from inland municipalities. Similar spatial patterns were described in a previous epidemiological investigation conducted at the same referral center between 2005 and 2015 (8), suggesting persistent structural challenges in the organization of leukemia care in the Amazon region. In this context, mortality from a potentially curable disease such as ALL reflects not only biological risk but also the performance and equity of the regional health system.

From a clinical perspective, most patients presented with advanced disease and high-risk features at diagnosis, including elevated leukocyte counts, anemia, thrombocytopenia, and a high proportion classified as high risk. This profile is consistent with reports from other low- and middle-income settings, where delays in diagnosis and referral contribute to increased disease burden at presentation and adversely affect outcomes (5,12,13). The predominance of B-cell ALL and the low frequency of atypical immunophenotypes suggest that the elevated mortality observed in this cohort is unlikely to be driven by uncommon biological subtypes, but rather by disease severity at presentation and contextual limitations in clinical management (8).

Comorbidities were frequent during follow-up, particularly infectious complications, underscoring their relevance in the clinical course of ALL in this setting (14,15). Multivariate analyses demonstrated that infectious comorbidities were not independently associated with sociodemographic characteristics, suggesting that these events are more closely related to treatment exposure, disease course, and healthcare trajectories than to baseline social factors (16,17). In contrast, non-infectious comorbidities were strongly associated with older age, reflecting the accumulation of chronic conditions over the life course, but were not independently associated with mortality (18,19).

Age at diagnosis emerged as the strongest determinant of mortality in this cohort. Adolescents, young adults, and patients aged 51–60 years exhibited significantly higher odds of death compared with younger children, consistent with extensive evidence demonstrating inferior outcomes outside the pediatric age range (20,21). These findings indicate that the survival gap described in high-income countries also persists in resource-limited contexts such as the Brazilian Amazon (8,22).

From a public health standpoint, the strong association between age at diagnosis and mortality highlights persistent gaps in the organization of leukemia care across the life course. Adolescents and young adults represent a transitional group that often falls between pediatric and adult oncology services, frequently experiencing delays in diagnosis, treatment discontinuity, and reduced access to structured supportive care (20,21). In geographically vulnerable settings such as the Brazilian Amazon, these challenges may be further amplified by the centralization of specialized services and long travel distances, contributing to avoidable mortality and reinforcing age-related inequities in outcomes (8,22).

Notably, the inverse association between infectious comorbidities and mortality should be interpreted within a health system context rather than as a protective biological effect. The occurrence of infectious complications likely reflects prolonged engagement with healthcare services, closer clinical surveillance, and timely access to supportive interventions, particularly in specialized referral centers (23,24). The concordance between regression models and survival analyses strengthens this interpretation. In contrast, non-infectious comorbidities were not associated with differences in overall survival (25,26), reinforcing the notion that these conditions primarily reflect baseline health status and age-related vulnerability rather than acting as direct drivers of mortality in ALL.

Collectively, these findings underscore that mortality patterns in ALL can serve as a sensitive indicator of health system performance and equity in access to specialized oncology care. Strengthening regional referral networks, improving early diagnosis in inland municipalities, enhancing continuity of care for adolescents and young adults, and expanding supportive management capacity in geographically remote areas are essential strategies to reduce avoidable mortality in vulnerable regions.

## LIMITATIONS

This study has limitations. Its retrospective design and reliance on secondary data may have resulted in incomplete information for selected variables, potentially affecting statistical power in some analyses. As a single-center study conducted at a tertiary referral hospital, the findings may not be fully generalizable; however, the institution captures the majority of ALL cases in the state of Amazonas. In addition, the lack of complete molecular and cytogenetic data limited more detailed risk stratification. Finally, the observational design precludes causal inferences, particularly regarding the associations between comorbidities and mortality, which should be interpreted as markers of clinical course and healthcare trajectories rather than direct effects.

## CONCLUSION

This study demonstrates that mortality from acute lymphoblastic leukemia in the Brazilian Amazon remains unacceptably high and is primarily associated with age at diagnosis and health system–related factors during treatment rather than baseline sociodemographic characteristics. Adolescents, young adults, and middle-aged patients exhibited disproportionately higher mortality, highlighting persistent gaps in the organization of leukemia care across the life course.

The substantial burden of comorbidities, particularly infectious complications, further underscores the importance of continuous access to specialized services and timely supportive management. In geographically vast and structurally vulnerable regions characterized by centralized high-complexity care, such as the Brazilian Amazon, mortality patterns in ALL reflect broader inequities in health system performance.

Strengthening referral networks, improving early diagnosis in inland municipalities, ensuring continuity of care for adolescents and young adults, and expanding supportive care capacity are essential strategies to reduce avoidable deaths. Addressing these structural determinants is critical to advancing equity in cancer outcomes in resource-limited and geographically vulnerable settings.

## DECLARATIONS

### Ethics approval and consent to participate

This study was approved by the Ethics Committee of the Fundação Hospitalar de Hematologia e Hemoterapia do Amazonas (HEMOAM) under approval number 4.982.395/2021. The requirement for written informed consent was waived by the Ethics Committee due to the retrospective design and the use of secondary data obtained from medical records, which posed minimal risk to participants. All procedures were conducted in accordance with the ethical principles of the Declaration of Helsinki and in compliance with Brazilian National Health Council Resolution No. 466/2012. Patient confidentiality and data anonymity were strictly maintained throughout the study.

### Consent for publication

Not applicable.

### Data availability

Due to ethical restrictions and the presence of potentially identifiable clinical information, the dataset generated and analyzed during the current study is not publicly available. De-identified data may be made available upon reasonable request to the corresponding author, subject to approval by the Ethics Committee of the Fundação Hospitalar de Hematologia e Hemoterapia do Amazonas (HEMOAM) and in accordance with institutional and national regulations governing the use of health data.

### Author contributions

IVS, FMG, FSAH, and AGC conceptualized the study. IVS, BSO, EYCO, JGSG, LPAC, JRFF, VGRS, JACN, FSP, JSM, MSB, FSS, and CLSC were responsible for data collection and investigation. IVS, FMG, MGAR, FSAH, and AGC performed the formal analysis. AGC acquired funding and supervised the project. IVS, BSO, and LPAC contributed to the methodology. FMG, FSAH, and AGC administered the project. IVS, FMG, and FSAH drafted the original manuscript. IVS, FMG, SVS, FSAH, and AGC critically revised the manuscript. All authors read and approved the final manuscript.

## Acknowledgments

The authors thank Fundação Hospitalar de Hematologia e Hemoterapia do Amazonas (HEMOAM) for granting access to institutional databases and the Rosa Blaya and RHC database teams for their support.

## SUPPLEMENTARY MATERIAL

**Supplementary Table 1.** Univariate logistic regression analysis of factors associated with infectious and non-infectious comorbidities among patients with acute lymphoblastic leukemia in Amazonas, Brazil, from 2016 to 2021.

**Supplementary Table 2.** Univariate logistic regression analysis of factors associated with mortality among patients with acute lymphoblastic leukemia in Amazonas, Brazil, from 2016 to 2021.

**Supplementary Figure 1.** Overall survival of patients with acute lymphoblastic leukemia according to non-infectious comorbidity status.

